# High-throughput metabolome comparison of cutaneous squamous cell carcinoma, basal cell carcinoma, and healthy skin with e-biopsy sampling

**DOI:** 10.1101/2024.03.06.24303646

**Authors:** Leetal Louie, Julia Wise, Ariel Berl, Ofir Shir-az, Vladimir Kravtsov, Zohar Yakhini, Avshalom Shalom, Alexander Golberg, Edward Vitkin

## Abstract

A standard histopathology-slides based diagnostics becomes a serious process bottleneck due to rising incidence rates of cutaneous squamous cell carcinoma (cSCC) and basal cell carcinoma (BCC). Leveraging tissue molecular information for diagnostics can be a beneficial alternative in certain cases. Sampling and processing of a constantly growing number of tumors can be enhanced with faster specimen collection methods together with high-throughput molecular identification approaches. Tumor specimens can be collected with electroporation-based biopsy (e-biopsy), a minimally invasive sampling collection tool with a proven ability, while mass spectrometry can be used for molecular identification.

The aim of this study was (i) to confirm the ability of e-biopsy technique to harvest metabolites, (ii) to obtain high-resolution metabolomic profiles of cSCC, BCC, and healthy skin tissues, and (iii) to perform a comparative analysis of the collected profiles.

Data, collected with e-biopsy coupled with ultra performance liquid chromatography and tandem mass spectrometry (UPLC-MS-MS), expands the current metabolomic profiles reported for cSCC, BCC, and healthy skin. Here we report measurements of 2325 small metabolites identified (301 with high confidence) in 13 tissue samples from 12 patients. Comparative analysis identified 34 significantly (p<0.05) differentially expressed high-confidence metabolites. Generally, we observed a greater number of metabolites with higher expression, in cSCC and in BCC compared to healthy tissues, belonging to the subclass amino acids, peptides, and analogues.

## Introduction

There is an observed rise in incidence rates of cutaneous squamous cell carcinoma (cSCC) and basal cell carcinoma (BCC)^1–5^. These cancers are commonly diagnosed but often not included in cancer registries due to their low mortality rate^4^, despite their impact on quality of life and the risks of premature mortality^2,6^. These trends bring attention to a potential increase in diagnostic wait times using the current gold standard method of tissue excision followed by histopathological examination^7^. Excision may also be problematic when subsequent treatment requires electrodesiccation and cautery^8^. In consideration of this, it is important to look for less invasive and time-efficient methods for distinguishing between healthy and cancerous skin, as well as between different types of skin cancers. Rapid methods would be beneficial considering the varying aggressiveness and likelihood of metastasis of cSCC compared to BCC^9^.

The potential of molecular profiling in diagnostics is evident given the observed capability to collect samples of molecular information from cSCC and BCC lesioned skin with simpler and faster methods than the current gold standard. Electroporation-based biopsy (e-biopsy) method leverages electric fields to induce permeabilization of cell membranes for harvesting molecular information. The analysis of molecular information obtained from specimens collected with this method has successfully differentiated between healthy and cancerous skin tissues^10–12^. Combined with high-throughput analytics that have demonstrated their ability to identify genes, proteins, lipids, and metabolites of cSCC and BCC^13–20^, e-biopsy is a promising direction for analysis of water-soluble molecules of skin cancers. E-biopsy broadens the spectrum of capabilities of handheld devices in cancer diagnostics and differs from tools such as the iKnife^21,22^ and MasSpec Pen^23,24^ which have been designed for intraoperative use and require real-time connection to a mass spectrometer.

Metabolomic analyses of cSCC and BCC have previously reported differences in cancerous tissues compared to healthy^14–16,18,19^. These reports have used a variety of analysis methods such as high-resolution magic angle spinning (HR-MAS) 1H nuclear magnetic resonance (NMR) spectroscopy^14^, liquid chromatography tandem mass spectrometry (LC-MS-MS)^15^, triple-quadrupole MS (QqQMS)^16^, ultraperformance LC coupled to a time-of-flight tandem MS (UPLC-TOF-MS/MS)^18^, and 1H NMR spectroscopy^19^, and have identified 9, 27, 27, 181, and 8 significant metabolites, respectively. These studies used both tissue^14,15,18,19^ and serum^16^ samples. Lacking here are promising minimally invasive specimen collection methods that can provide inputs into these high-throughput analyses for metabolite identification.

Here we present and perform the first comparative analysis of high-throughput metabolomic profiling of cSCC, BCC, and healthy skin tissues sampled with the e-biopsy method. This study contributes to the recently discovered capabilities of e-biopsy in transcriptomics, proteomics, and lipidomics^10–13,20^.

## Materials and Methods

### Human patients

**Table 1** contains patient sex, age, and tumor type. This study was approved by the Meir Medical Center IRB, number MMC-19-0230. All patients gave consent for participation and performance of genetic analysis of their sample tissue.

**Table 1.** Patient sex, age, and tumor type.

| Age range | Sex | Tumor type |
| --- | --- | --- |
| Below 70 | Male | cSCC |
|  | Female | BCC |
|  | Female | Healthy |
| 70-80 | Male | cSCC |
|  | Male | BCC |
|  | Female | BCC |
|  | Female | Healthy |
| Above 80 | Female | cSCC |
|  | Male | cSCC |
|  | Female | BCC |
|  | Male | BCC |
|  | Female | BCC |

### Sample collection

Ten tissue samples were collected from 10 patients undergoing surgical excision of cSCC or BCC between March 2020 and March 2022 at Meir Medical Center, Israel. Additionally, 3 healthy tissue samples were collected from 2 patients undergoing blepharoplasty. The diameter of excised tissue was at least 1cm. Between 10-20 minutes after surgery, a total of 13 fresh tissue samples underwent e-biopsy extraction methods. Following this, UPLC-MS-MS and differential expression analyses were performed. The workflow of the e-biopsy and subsequent analysis are summarized in **Fig. 1**.

**Fig. 1.**
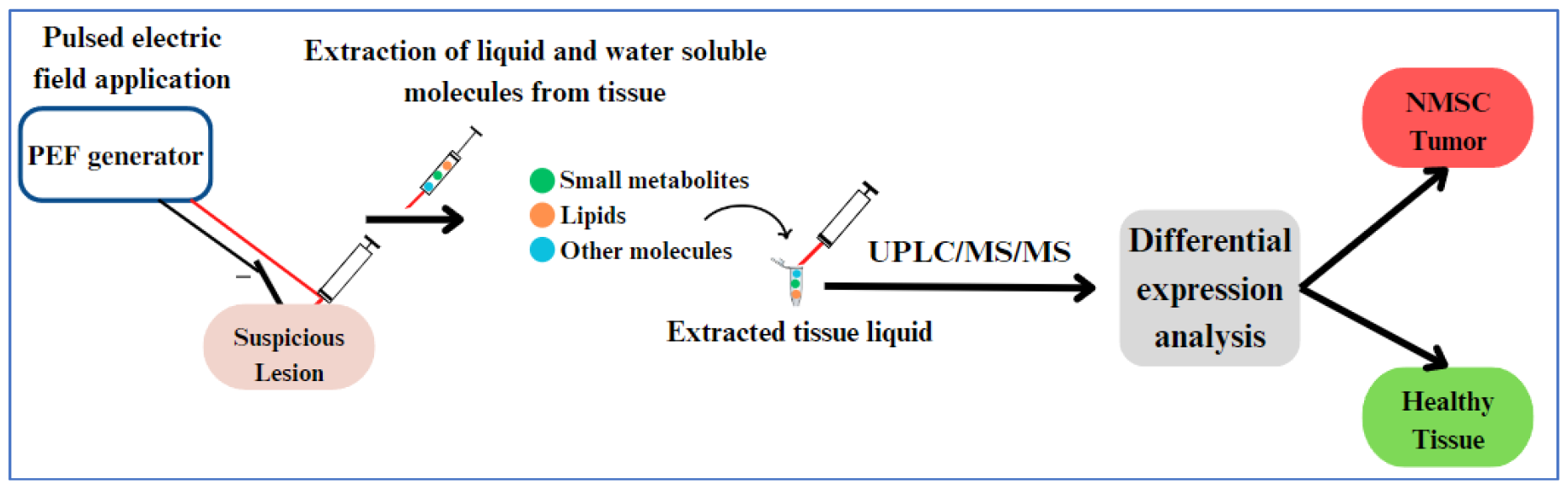
Study workflow: A pulsed electric field (PEF) is delivered to a tissue sample for extraction of water-soluble molecules and sample extracts are stored in 1.5 ml tubes until ready for UPLC-MS-MS analysis. Data from UPLC-MS-MS is used for differential expression analysis.

A standard 30-G insulin syringe with a needle is inserted in the sampling location and the ground electrode (custom made with a 3mm diameter) is positioned approximately 5 mm apart on the skin surface, without penetration. A pulsed electric field (PEF) is delivered through the sampling needle and then a vacuum is applied to the needle to drive the released cellular content into it and the syringe. PEF settings are a combination of high-voltage short pulses and low-voltage long pulses^10,25^: 40 pulses, 1000 V, 40 μs, 4 Hz, and 40 pulses, 50 V, 5 ms, 4 Hz. The syringe used to collect liquids from tissues is 1.5 ml and the vacuum on the needle is applied manually. Immediately following, liquids are transferred to 1.5ml tubes that contained 100μl double distilled water and placed in -20□. A custom-made high voltage pulsed electric field generator is used for PEF application^26^.

Samples were stored until shipped to Beijing Genomics Institute for analysis.

### UPLC-MS-MS Analysis

UPLC-MS-MS analysis was performed by Beijing Genomics Institute. An ACQUITY UPLC BEH C18 column and UPLC BEH Amide column (both 1.7 μm, 2.1 mm x 100 mm, Waters, USA) in both positive and negative modes were used for chromatographic separation (4 analyses in total). Water 2D UPLC (Waters, USA) and tandem Q Exactive high resolution mass spectrometer (Thermo Fisher Scientific, USA) with a heated electrospray ionization (HESI) source were used for this analysis. Xcalibur 2.3 software was used. Data gathered included metabolite ID, ID reliability level (graded level 1 to 4, with 1 and 2 being the most accurately identified metabolites used for subsequent differential screening), and metabolite intensity (**Table S1, Table S2, Table S3, Table S4**). This data was used for the analysis of differential metabolite abundance. Detailed UPLC-MS-MS information and methods can be found in the **Supplementary methods**.

### Differential metabolite screening

Differential metabolite screening was performed on all metabolite intensities measured by UPLC-MS-MS (**Table S1, Table S2, Table S3 and Table S4**). Student’s t-test and fold change between average metabolite intensities was performed for each metabolite in each comparison pair (cSCC vs. healthy, BCC vs. healthy, and cSCC vs. BCC). Resulting p-values and fold change values were used for overabundance analysis and volcano analysis (**Fig. 2 and Fig. 3**).

**Fig. 2.**
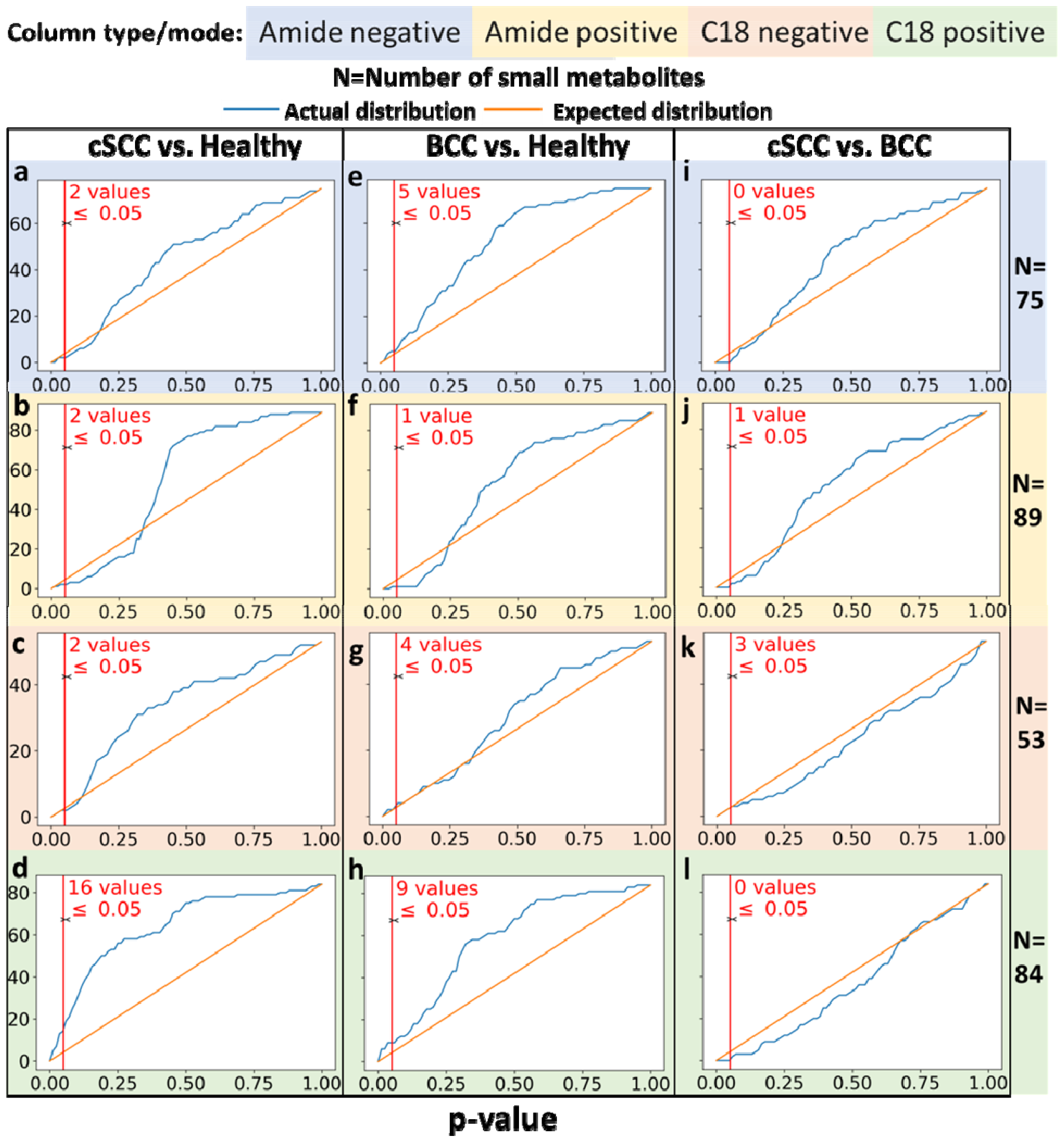
**a-l**. Overabundance plots comparing the distribution of metabolites differential expression (both over- and under-expression) p-values between control (normal skin tissue), cSCC, BCC tumor samples. Results from 2 chromatography column types and mode is shown by the background color. A total of 13 samples, and 301 metabolites extracted by e-biopsy were analyzed. **a-d**. cSCC vs. healthy **e-h**. BCC vs. healthy and **i-l**. cSCC vs. BCC.

**Fig. 3.**
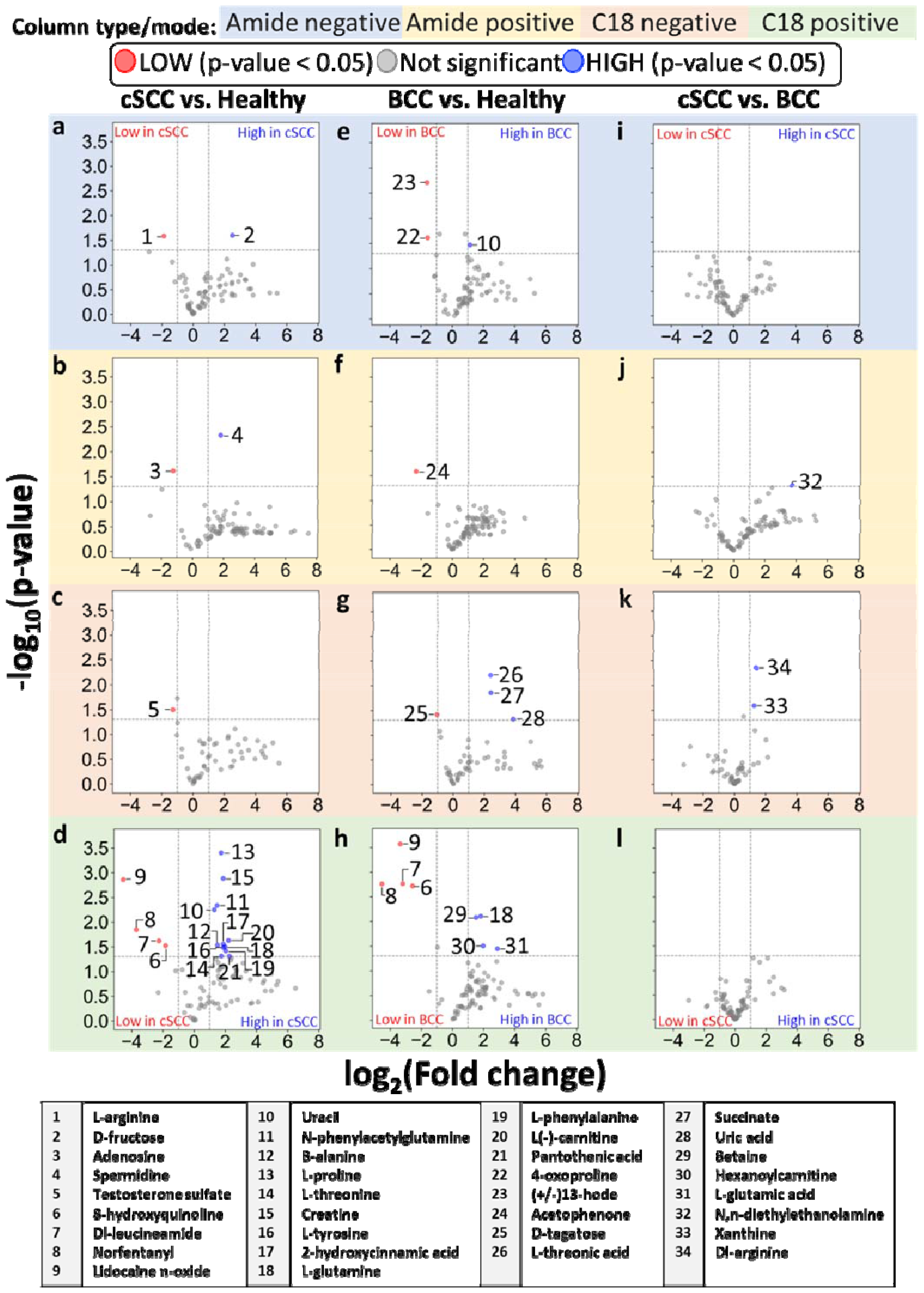
**a-l**. Volcano plots showing the fold-change difference of metabolite intensities. **a-d**. cSCC vs. Healthy. **b**. BCC vs. Healthy. **c**. cSCC vs. BCC. Numbered datapoints correspond to metabolite names found in the table below the plots. Fold-change and p-value data for the metabolites identified in the table can be found in **Table 2, Table S5, Table S6, and Table S7**. High-resolution plots can be found in **Figs. S1-12**.

#### Statistical overabundance analysis

Overabundance analysis verifies that compounds have different abundance levels when comparing two classes of samples by comparing the actual and expected distribution of p-values^27^. This analysis is used in analyses with multiple comparisons and explores internal data variability. The analysis relies only on the number of compounds (i.e., metabolites) and their observed p-values (obtained here from the Student’s t-test). P-value correction was calculated with the Bonferroni-Hochberg approach. FDR was calculated as the ratio of expected and received values below an uncorrected p-value of interest. The distribution of the expected p-values was generated from a null model assuming the same number of compounds (**Fig. 2**).

**Table 2.** Differentially expressed metabolites with associated fold change values and p-values. Columns contain the metabolites (in bold) identified as significant by the pairwise comparison groups cSCC vs. Healthy, BCC vs. Healthy and cSCC vs. BCC. An additional column is added for metabolites found in both cSCC and BCC compared to Healthy. **Reading example:** Dl-arginine ratio of its average intensity in cSCC tissues to average intensity in BCC tissues is 2.71. This comparison has a Student T-test p-value of 0.005.

| **cSCC vs. Healthy** Fold-change (p-value) | **BCC vs. Healthy** Fold-change (p-value) | **cSCC vs. BCC** Fold-change (p-value) |
| --- | --- | --- |
| **L-glutamine** 3.97 (0.03) | **L-glutamine** 3.54 (0.008) | **DL-arginine** 2.71 (0.005) |
| **DL-leucineamide** 0.21 (0.02) | **DL-leucineamide** 0.11 (0.002) | **Xanthine** 2.40 (0.03) |
| **Norfentanyl** 0.08 (0.01) | **Norfentanyl** 0.04 (0.002) | **N,n-diethylethanolamine** 13.15 (0.05) |
| **Lidocaine n-oxide** 0.04 (0.001) | **Lidocaine n-oxide** 0.1 (0.0003) |  |
| **8-hydroxyquinoline** 0.28 (0.03) | **8-hydroxyquinoline** 0.17 (0.002) |  |
| **Uracil** 2.46 (0.006) | **Uracil** 2.20 (0.03) |  |
| **Testosterone sulfate** 0.41(0.03) | **Succinate** 5.38 (0.01) |  |
| **L(-)-carnitine** 4.60 (0.02) | **L-threonic acid** 5.31 (0.006) |  |
| **B-alanine** 2.83 (0.03) | **D-tagatose** 0.49 (0.04) |  |
| **L-threonine** 3.37 (0.05) | **Uric acid** 14.58 (0.05) |  |
| **Creatine** 3.66 (0.001) | **L-glutamic acid** 7.46 (0.04) |  |
| **L-proline** 3.34 (0.0004) | **Betaine** 2.87 (0.008) |  |
| **2-hydroxycinnamic acid** 3.67 (0.03) | **Hexanoylcarnitine** 3.96 (0.03) |  |
| **L-tyrosine** 3.66 (0.03) | **(+/-)13-hode** 0.33 (0.002) |  |
| **L-phenylalanine** 4.11 (0.04) | **4-oxoproline** 0.34 (0.02) |  |
| **Pantothenic acid** 4.79 (0.05) | **Acetophenone** 0.2 (0.03) |  |
| **N-phenylacetylglutamine** 2.79 (0.005) |  |  |
| **L-arginine** 0.27 (0.03) |  |  |
| **D-fructose** 5.75 (0.03) |  |  |
| **Spermidine** 3.54 (0.005) |  |  |
| **Adenosine** 0.42 (0.02) |  |  |

#### Volcano plot analysis

Volcano plot depicts the magnitude of fold change of metabolites and differential significance between two analyzed populations. Differentially expressed metabolites were defined for this analysis as those with a *-log10(p-value)>1.3* (i.e. *p-value<0.05*) and those with *-1<log2(fold-change)<1*. Fold change was calculated using the average of intensity values for each comparison group i.e., *fold-change(metabolite) = avg(grp1) / avg(grp2)*. The data was then filtered to include only the compounds with high reliability score (levels 1 and 2) (**Fig 3)**. The data of the most interesting compounds were organized into tables to showcase their associated p-values and fold change values (**Table 2, Tables S5, Table S6 and Table S7**).

## Results

A total of 2325 metabolites were identified across 2 different columns in 2 modes (**Methods**) and then filtered to 301 eligible for differential expression according to the measurement reliability level, omitting metabolites with level 3 and 4. Eligible metabolites were used for the following groupwise comparisons: cSCC vs. Healthy, BCC vs. Healthy, and cSCC vs. BCC. Overabundance plots (**Fig. 2**) present analysis results, and the number of metabolites with Student T-test p-value below 0.05 is highlighted in red. Volcano plots (**Fig. 3**) show relative group affinity of each metabolite, highlighting significantly over- and under-expressed metabolites. The major findings are summarized in **Table 2**.

### cSCC vs. Healthy

Across all column types and modes, the overabundance analysis of cSCC compared to healthy skin revealed a total of 6 metabolites with Student T-test p-values below 0.01 (FDR=0.5) and 22 with p-values below 0.05 (including 3 after correction, FDR=0.68, **Fig. 2 a-d**), with 14 significantly (fold change>2) higher in cSCC and 7 significantly lower in cSCC (**Fig. 3 a-d)**. The amide column in negative mode (**Fig. 3a**) identified L-arginine at lower intensity and D-fructose at higher intensity in cSCC. The amide column in positive mode (**Fig. 3b**) identified adenosine at lower intensity and spermidine at higher intensity in cSCC. The C18 column in negative mode (**Fig. 3c**) identified testosterone sulfate at lower intensity in cSCC. The C18 column in positive mode (**Fig. 3d**) identified 8-hydroxyquinoline, DI-leucineamide, norfentanyl, and lidocaine n-oxide at lower intensity, and uracil, n-phenylacetylglutamine, B-alanine, L-proline, L-threonine, L-tyrosine, 2-hydroxycinnamic acid, L-glutamine, L-phenylalanine, L(-)-carnitine, pantothenic acid and creatine at higher intensity in cSCC. Moreover, l-proline was identified with p-value=4e-4 (corrected p-value 0.03). High-resolution volcano plot of this comparison can be viewed in the supplementary material (**Fig. S1, Fig. S2, Fig. S3, and Fig. S4**). All associated p-values and fold change values for the metabolites listed above can be found in **Table 2** and **Table S5**.

### BCC vs. Healthy

Across all column types and modes, the overabundance analysis of BCC compared to healthy skin revealed a total of 8 metabolites with Student T-test p-values below 0.01 (FDR=0.38) and 19 with p-values below 0.05 (FDR=0.79), including 4 after Benjamini-Hochberg correction (**Fig. 2 e-h**), with 8 significantly (fold change>2) higher in BCC and 8 significantly lower in cSCC (**Fig. 3 e-h)**. The amide column in negative mode (**Fig. 3e**) identified 4-oxoproline and (+/-)13-hode at lower intensity and uracil at higher intensity in BCC. The amide column in positive mode (**Fig. 3f**) identified acetophenone at lower intensity in BCC. The C18 column in negative mode (**Fig. 3g**) identified D-tagatose at lower intensity and L-threonic acid, succinate, and uric acid at higher intensity in BCC. The C18 column in positive mode (**Fig. 3h**) identified 8-hydroxyquinoline, DI-leucineamide, norfentanyl, and lidocaine n-oxide at lower intensity, and L-glutamine, betaine, hexanoylcarnitine, and L-glutamic acid at higher intensity in BCC. Moreover, Dl-leucineamide was identified with p-value=2e-3 (corrected p-value 0.04). High-resolution volcano plot of this comparison can be viewed in the supplementary material (**Fig. S5, Fig. S6, Fig. S7, and Fig. S8**). All associated p and fold change values for the metabolites listed above can be found in **Table 2** and **Table S6**.

### cSCC vs. BCC

Across all column types and modes, the overabundance analysis of cSCC compared to BCC revealed a 1 metabolite (Dl-arginine) with Student T-test p-values below 0.01 and 4 metabolites with p-values below 0.05 (FDR>1, **Fig. 2i-l**), with 3 significantly (fold change>2) higher in cSCC and none significantly lower (**Fig. 3i-l)**. The amide column in negative mode (**Fig. 3i**) did not identify differentially expressed metabolites. The amide column in positive mode (**Fig. 3j**) identified N,n-diethylethanolamine at higher intensity in cSCC. The C18 column in negative mode (**Fig. 3k**) identified xanthine and DI-arginine at higher intensity in cSCC. The C18 column in positive mode (**Fig. 3l**) did not identify differentially expressed metabolites. High-resolution volcano plot of this comparison can be viewed in the supplementary material (**Fig. S9, Fig. S10, Fig. S11, and Fig. S12**). All associated p and fold change values for the metabolites listed above can be found in **Table 2** and **Table S7**.

## Discussion

A comparative high-throughput analysis of metabolomic profiles was performed on cSCC, BCC, and healthy skin tissues sampled with e-biopsy technique. The results suggest a difference in metabolomic profiles between cancer and healthy tissues. The most notable trends in the data are the abundance of metabolites in the subclass of amino acids, peptides, and analogues. Of the total 34 identified differentially expressed metabolites, 14 fall into this subclass. Of the 14 in this subclass, 10 were found to be present in higher quantities in the cancerous tissues compared to healthy tissues and 1 higher in cSCC compared to BCC. Additionally, the 2 significant metabolites of the subclass carbohydrates and carbohydrate conjugates were both found to be higher in the cancerous tissues compared to healthy. There do not appear to be comparison-group wide trends for the remaining significant compounds.

To the best of our knowledge, no previous metabolomic profiling studies using e-biopsy molecular harvesting from cSCC and BCC were performed. Previous studies comparing cSCC to healthy skin have used QqQMS^16^, UPLC-TOF-MS/MS and nLC-MS/MS^18^, and 1H NMR spectroscopy^19^. Previous studies comparing BCC to healthy skin tissue have used high-resolution magic angle spinning 1H NMR^14^ and LC-MS-MS^15^. An advantage of the e-biopsy method is that sample preparation does not require overnight storage as with some of these previously noted methods^16,18,19^.

Our study contributes to the growing body of knowledge of molecular markers of cSCC and BCC, helping to pave the way for molecular diagnostics with potential to outperform the current methods. E-biopsy coupled with UPLC-MS-MS was able to reliably identify 301 different metabolites, with similarities to previous metabolomic studies of cSCC and BCC. As with our study, a previous study found phenylalanine, fructose and tyrosine increased in cSCC versus healthy^16^. This study also found higher levels of glutamic acid in cSCC versus healthy^16^, however we found glutamic acid differentially expressed in BCC versus healthy samples only. Like in our study, there are previous reports of increased uracil in BCC versus healthy samples^28^, however, we also found higher levels of uracil in cSCC versus healthy. Pantothenic acid and betaine were previously reported to be lower in BCC compared to healthy skin^15^, whereas we found higher betaine in BCC and higher pantothenic acid in cSCC. Unlike previous research that identified lower creatine in BCC compared to healthy skin^14^, we observed higher creatine in comparison of cSCC to healthy skin tissue. A previous study of cSCC compared to healthy skin found higher levels of creatine, tyrosine, and glutamine^19^, which is consistent with our results. Again, consistent with our results, another comparison of cSCC to healthy found threonine, creatine, proline, phenylalanine, pantothenic acid, spermidine, uracil, and glutamine higher in cSCC compared to healthy^18^. This study also found higher adenosine present in cSCC compared to healthy skin^18^, whereas we observed it in lower levels in cSCC. The same study found that arginine was higher in cSCC compared to healthy^18^, but our study observed arginine lower in cSCC compared healthy though higher in cSCC compared to BCC. The difference in the arginine measurements may result from different enantiomers, but this is not clear from the data previously reported^18^. Other metabolites identified by UPLC-TOF-MS/MS and nLC-MS/MS that were also identified here are glutamic acid, betaine, and hexanoylcarnitine^18^. These 3 metabolites however, were not identified as significant in the comparison of cSCC to healthy, rather they were significant in BCC compared to healthy. Lastly, xanthine was previously found significantly higher in cSCC compared to healthy skin^18^, whereas we found it higher in cSCC compared to BCC. The similarities observed demonstrate that the results obtained from e-biopsy-produced samples are consistent with other methods for metabolite profiling.

Several potentially interesting differentially expressed metabolites were identified in our study. Cinnamic acid, whose derivative is hydroxycinnamic acid, has been observed to reduce proliferation of cells in glioblastoma, melanoma, prostate, and lung cancers^29^. Additionally, cinnamic acid and its derivatives have been shown to trigger apoptosis in melanoma cells^30^. As with 2-hydroxycinnamic acid, phenylalanine and tyrosine which are precursors to cinnamic acid^31^ were also found in higher quantities in cSCC samples. Phenylalanine and tyrosine have also both been examined as biomarkers of skin cancer^32^. B-alanine, found higher in cSCC, is involved in mechanisms of itch and pain^33^ which are symptoms associated with cSCC^34^. Creatine, which we found at higher levels in cSCC, is created in the generation of ATP^35^ that provides energy for cells. SCC and BCC have been found to have higher total activity of certain creatine kinase isozymes compared to normal human skin^35^. L-proline, which we found higher in cSCC, was reported to suppress actinic skin cell damage after solar stimulated UV radiation^36^, possibly suggesting that it is produced in abundance in cSCC as a defense mechanism. Follow-up studies with in-depth analysis of specific metabolites and a larger patient cohort should be performed for validation of these findings.

Limitations of this study include a sample size not representative enough for drawings conclusions about metabolite behavior. Comparison with previous studies exemplifies the benefit of larger samples for identifying trends in skin cancer lipids^37,38^, thus an increased sample size can improve confidence in findings of differential expression metabolomic analysis and can reduce the amount of falsely detected signals. Additionally, this study was performed on *ex-vivo* samples extracted by standard excision biopsy which might produce different results than an *in-vivo* analysis. Lastly, patient inclusion criteria were not very strict, potentially overlooking patient lifestyle information that could provide useful information such as levels of sun exposure.

The early-stage results of the e-biopsy sampling technique show promise of its potential development as a handheld device that limits the need for tissue resection during biopsy. Its ability to be used as a standalone device provides advantages over tools such as the iKnife^21,22^ and MasSpec Pen^23,24^ in primary care settings. In contrast to other needle biopsy techniques such as fine-needle aspiration and core needle biopsy, needle size is not a limiting factor^39,40^, and a more invasive larger needle diameter is not needed for greater accuracy^41,42^. Conversely, e-biopsy can provide valuable site-specific information by its ability to sample areas larger than the needle diameter ^12^ which provides opportunities for greater understanding of tumor complexities and mapping of tumor heterogeneity^11^.

## Conclusion

This study contributes to the understanding of cSCC and BCC molecular mechanisms. We report high-throughput metabolomics profiles of cSCC, BCC and healthy skin, together with a comparative analysis between the 3 tissue types. A total of 2325 metabolites were identified, among them 301 with high confidence. In that group 34 metabolites were found to be differentially expressed. The differentially expressed metabolites were from several subclasses (e.g. amino acids, peptides, and analogues, and carbohydrates and carbohydrate conjugates). Classification details are found in the supplementary material (**Table S1, Table S2, Table S3, and Table S4**). Overall trends in the data indicate a greater number of amino acids, peptides, and analogues with higher expression in both cSCC and in BCC compared to healthy skin tissue. This study demonstrates the opportunity in high-throughput metabolomic profiling of e-biopsy-gathered samples and its potential for enhancing skin caner diagnostic methods.

## Data Availability

All the data that supports the findings of this study are available in the supplementary materials.

## Abbreviations

BCC: basal cell carcinoma
cSCC: cutaneous squamous cell carcinoma
e-biopsy: electroporation-based biopsy
UPLC-MS-MS: ultra performance liquid chromatography and tandem mass spectrometry

## Data accessibility

All the data that supports the findings of this study are available in the supplementary materials.

## Author contributions

LL – conceptualization, experiments, metabolite sampling and analysis, data preparation and analysis, bioinformatics, manuscript drafting

JW – experiments, metabolite sampling and analysis

AB– experiments, samples collection, pathology, clinics, manuscript review

OS– experiments, samples collection, pathology, clinics

VK – pathology

ZY – conceptualization, data analysis

AS – conceptualization, critical manuscript review

AG– conceptualization, experiments, data analysis, manuscript drafting

EV – conceptualization, bioinformatics, manuscript drafting and approval All authors contributed to the manuscript review.

## Acknowledgements

The authors thank Beijing Genomic Institute for metabolomics services.

