## Supplementary methods for "High-throughput metabolome comparison of cutaneous squamous cell carcinoma, basal cell carcinoma, and healthy skin with e-biopsy sampling"

**UPLC-MS-MS Analysis**

 The following sections are presented as reported by BGI Group.

***Main Instruments and Reagents***

Ultra Performance Liquid Chromatography:(Waters 2D UPLC, Waters, USA)

High Resolution mass spectrometer:(Q Exactive, Thermo Fisher Scientific, USA)

Chromatographic Column: ACQUITY UPLC BEH C18 (1.7 μm,2.1*100 mm,Waters, USA)

ACQUITY UPLC BEH Amide (1.7 μm,2.1*100 mm, Waters, USA)

Low Temperature High Speed Centrifuge:(Centrifuge 5430, Eppendorf)

Vortex:(QL-901, Kylin-bell Lab Instruments Co., Ltd.,China)

Ultrapure Water Systems:(Milli-Q Integral,Millipore Corporation,USA)

Vacuum Concentrator:(Maxi Vacbeta,GENE COMPANY)

Internal standard mix (IS) contains: L-Leucine-d3,L-PHENYLALANINE (13C9, 99%),L- Tryptophan-d5,Progesterone-2,3,4-13C3.

MS-grade methanol (A454-4) and acetonitrile (A996-4) were purchased from Thermo Fisher Scientific (USA). Formic acid was purchased from DIMKA (50144-50ml, USA) and ammonium formate (17843-250G,Honeywell Fluka, USA) was obtained from Fluka. Ultrapure water was filtered through the Milli-Q system.

***Small Metabolites Extraction***

Metabolite extraction was primarily performed according to previously reported methods [6][7]. In short, 100 μL samples were extracted by directly adding 300 μL of precooled methanol and acetonitrile (2:1, v/v), internal standards mix 1 (IS1) and internal standards mix 2 (IS2) were added for quality control of sample preparation. Samples were vortexed for 1 minute and incubated at -20 °C for 2 hours. Following this, samples were centrifuged for 20 min at 4000 rpm, and the supernatant was then transferred for vacuum freeze drying. The metabolites were resuspended in 150 μL of 50% methanol and centrifuged for 30 min at 4000 rpm, and the supernatants were transferred to autosampler vials for LC-MS analysis. A quality control (QC) sample was prepared by pooling the same volume of each sample to evaluate the reproducibility of the whole LC-MS analysis.

***UPLC-MS Analysis***

*Liquid chromatography conditions – amide column:*

The samples were analyzed on a Waters 2D UPLC (Waters, USA), coupled to a Q-Exactive mass spectrometer (Thermo Fisher Scientific, USA) with a heated electrospray ionization (HESI) source and controlled by the Xcalibur 2.3 software program (Thermo Fisher Scientific, Waltham, MA, USA). Chromatographic separation was performed on a Waters ACQUITY UPLC BEH C18 column (1.7 μm, 2.1 mm × 100 mm, Waters, USA), and the column temperature was maintained at 45 °C. The mobile phase consisted of 0.1% formic acid (A) and acetonitrile (B) in the positive mode, and in the negative mode, the mobile phase consisted of 10 mM ammonium formate (A) and acetonitrile (B). The gradient conditions were as follows: 0-1 min, 2% B; 1-9 min, 2%-98% B; 9-12 min, 98% B; 12-12.1 min, 98% B to 2% B; and 12.1-15min, 2% B. The flow rate was 0.35 mL/min and the injection volume was 5 μL.

*Liquid chromatography conditions – C18 column:*

The intracellular metabolite extracts were analyzed on a Waters 2D UPLC (Waters, USA), coupled to a Q-Exactive mass spectrometer (Thermo Fisher Scientific, USA) with a heated electrospray ionization (HESI) source and controlled by the Xcalibur 2.3 software program (Thermo Fisher Scientific, Waltham, MA, USA). Chromatographic separation was performed on a Waters ACQUITY UPLC BEH Amide column (1.7 μm, 2.1 mm × 100 mm, Waters, USA), and the column temperature was maintained at 30 °C. In positive mode, the mobile phase A consisted of 0.1% formic acid, 10 mM ammonium formate and 95% acetonitrile, the mobile phase B consisted of 0.1% formic acid, 10 mM ammonium formate and 50% acetonitrile. In negative mode, the mobile phase A consisted of 10 mM ammonium formate and 95% acetonitrile, the mobile phase B consisted of 10 mM ammonium formate and 50% acetonitrile. The gradient conditions were as follows: 0-0.5 min, 2% B; 0.5-12 min, 2%-50% B;

12-14 min, 98% B; 14-16 min, 98% B; 16-16.1 min, 98%-2% B and 16.1-18min, 2% B. The flow rate was 0.35 mL/min and the injection volume was 2 μL.

Positive ion mode, which refers to the positive ions of H+, NH4+, Na+, K+, etc. when the substance is ionized at the ion source.

Negative ion mode, which refers to the addition of ions to -H, +Cl plasma when the ion is ionized at the ion source.

*Mass spectrometry conditions:*

The mass spectrometric settings for positive/negative ionization modes were as follows: spray voltage, 3.8/−3.2 kV; sheath gas flow rate, 40 arbitrary units (arb); aux gas flow rate, 10 arb; aux gas heater temperature, 350 °C; capillary temperature, 320 °C. The full scan range was 70–1050 m/z with a resolution of 70000, and the automatic gain control (AGC) target for MS acquisitions was set to 3e6 with a maximum ion injection time of 100 ms. Top 3 precursors were selected for subsequent MSMS fragmentation with a maximum ion injection time of 50 ms and resolution of 17500, the AGC was 1e5. The stepped normalized collision energy was set to 15, 30, and 45 eV for the amide column, and 20, 40 and 60 eV for the C18 column.

***Data preprocessing***

The mass spectrometry raw data (raw file) collected by LC-MS/MS was imported into Compound Discoverer 3.1 (Thermo Fisher Scientific, USA) for data processing, including: peak extraction, retention time correction within and between groups, additive ion pooling. , missing value filling, background peak labeling, and metabolite identification, and finally information on compound molecular weight, retention time, peak area, and identification results were exported. The identification of metabolites is a combined result of BMDB database, mzCloud and ChemSpider (HMDB, KEGG, LipidMaps) databases.Main parameters of metabolite identification:Precursor Mass Tolerance<5

ppm,Fragment Mass Tolerance<10 ppm,RT Tolerance<0.2 min.

Compound Discoverer 3.1 results were exported to metaX for data preprocessing which included normalization with probabilistic quotient normalization (PQN), quality control-based robust LOESS signal correction, and calculation of the coefficient of variation (CV) of relative peak areas in all QC samples. Compounds with a CV >30% were filtered out.

There are 5 levels of confidence in compound identification results; Level 1: Substances that can be accurately determined based on the standard database and experimental data. Level 2: Substances whose structural formula matches the standards database. Level 3: Substances whose structural part can be matched with the standards database but need further verification. Level 4: Substances that match the first mass. Level 5: No substance is matched in the database, that is, there is no identification result.

***Data quality control***

The data quality was evaluated by the repeatability of QC sample detection. Quality control consisted of overlapping QC chromatograms to ensure signal stability through minimal fluctuation of retention time and peak response intensity. Principal component analysis (PCA) of log2 transformed data standardized with pareto scaling was used to observe separation trends between sample groups, identify abnormal points, and variation between and within groups. In this way, the overall distribution and stability of QC samples and all samples can be observed. Lastly, the differences in the number and peak area of lipid molecules underwent QC by examining the ratio of lipid molecules whose CV of relative peak area is less than or equal to 30% in QC samples to the number of all detected compounds. Data was qualified in the ratio was greater or equal to 60%.
