## Supplementary material for "High-throughput metabolome comparison of cutaneous squamous cell carcinoma, basal cell carcinoma, and healthy skin with e-biopsy sampling"

**Supplementary Figures’ Legends**

**Figure S1.** cSCC vs. healthy volcano analysis of the amide negative column output showing the fold change difference of metabolite intensity. Corresponding fold change and p-values are found in **Table** **S5.**

**Figure S2.** cSCC vs. healthy volcano analysis of the amide positive column output showing the fold change difference of metabolite intensity. Corresponding fold change and p-values are found in **Table S5**.

**Figure S3.** cSCC vs. healthy volcano analysis of the C18 negative column output showing the fold change difference of metabolite intensity. Corresponding fold change and p-values are found in **Table S5.**

**Figure S4.** cSCC vs. healthy volcano analysis of the C18 positive column output showing the fold change difference of metabolite intensity. Corresponding fold change and p-values are found in **Table S5**.

**Figure S5.** BCC vs. healthy volcano analysis of the amide negative column output showing the fold change difference of metabolite intensity. Corresponding fold change and p-values are found in **Table S6**.

**Figure S6.** BCC vs. healthy volcano analysis of the amide positive column output showing the fold change difference of metabolite intensity. Corresponding fold change and p-values are found in **Table S6**.

**Figure S7.** BCC vs. healthy volcano analysis of the C18 negative column output showing the fold change difference of metabolite intensity. Corresponding fold change and p-values are found in **Table S6**.

**Figure S8.** BCC vs. healthy volcano analysis of the C18 positive column output showing the fold change difference of metabolite intensity. Corresponding fold change and p-values are found in **Table S6**.

**Figure S9.** cSCC vs. BCC volcano analysis of the amide negative column output showing the fold change difference of metabolite intensity.

**Figure S10.** cSCC vs. BCC volcano analysis of the amide positive column output showing the fold change difference of metabolite intensity. Corresponding fold change and p-values are found in **Table S7.**

**Figure S11.** cSCC vs. BCC volcano analysis of the C18 negative column output showing the fold change difference of metabolite intensity. Corresponding fold change and p-values are found in **Table S7**.

**Figure S12.** cSCC vs. BCC volcano analysis of the C18 positive column output showing the fold change difference of metabolite intensity.

**Supplementary Tables’ Legends**

**Table S1.** List of all metabolites identified with a C18 column in positive mode.

**Table S2.** List of all metabolites identified with a C18 column in negative mode.

**Table S3.** List of all metabolites identified with an amide column in positive mode.

**Table S4.** List of all metabolites identified with an amide column in negative mode.

**Table S5.** cSCC vs. healthy differentially expressed metabolites.

**Table S6.** BCC vs. healthy differentially expressed metabolites.

**Table S7.** cSCC vs. BCC differentially expressed metabolites.
