## Supplementary figures and images for "High-throughput metabolome comparison of cutaneous squamous cell carcinoma, basal cell carcinoma, and healthy skin with e-biopsy sampling"

### Fig. S1

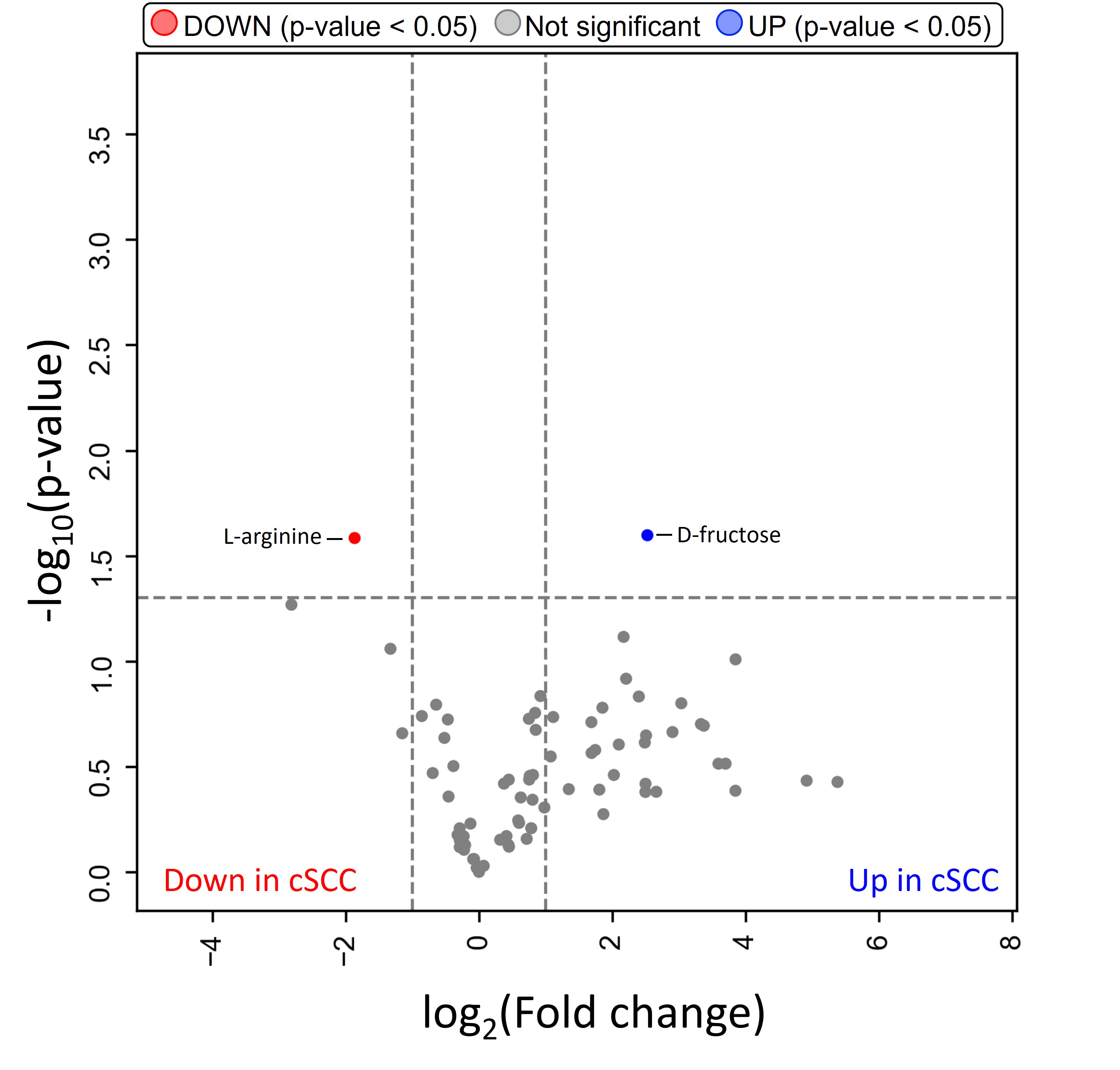

### Fig. S2

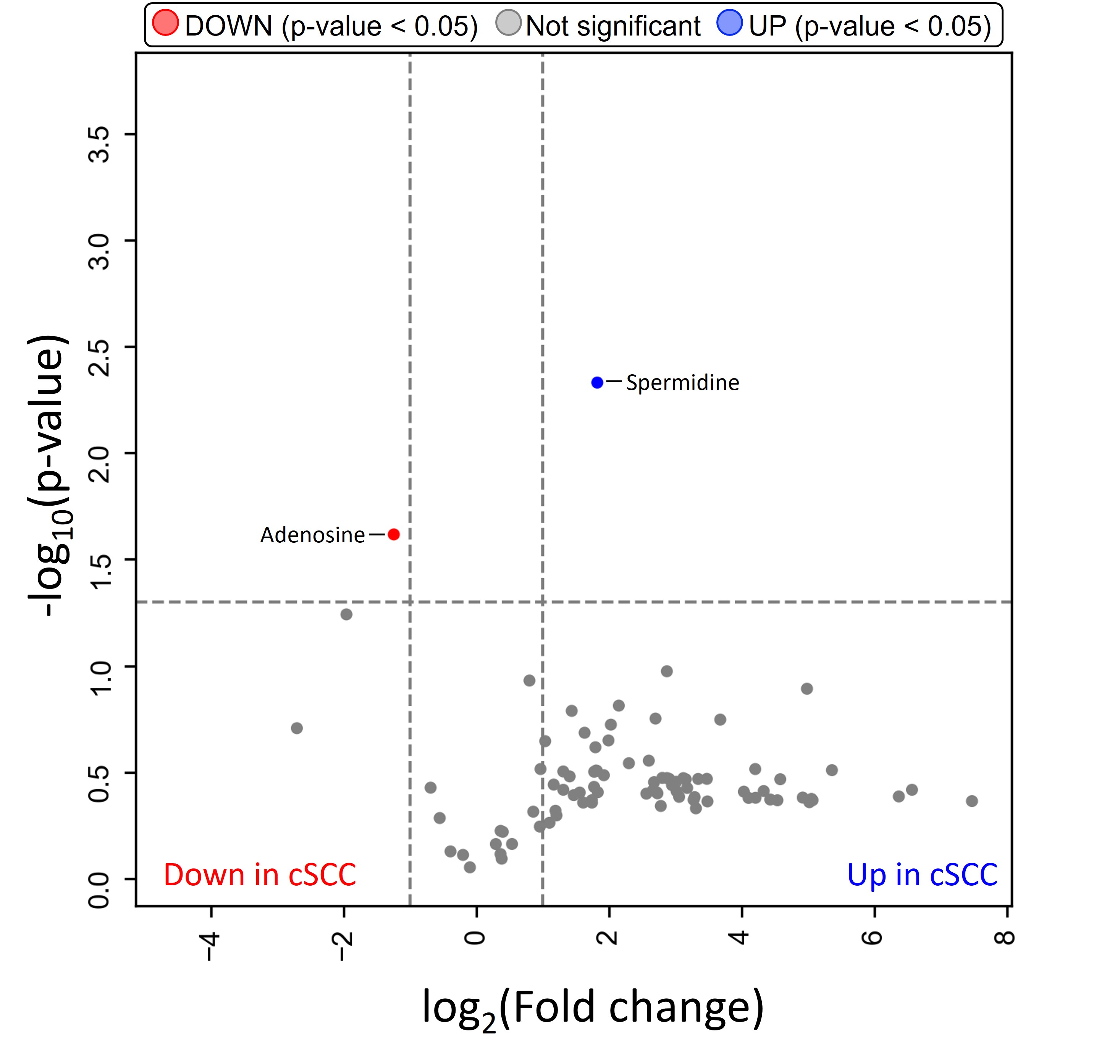

### Fig. S3

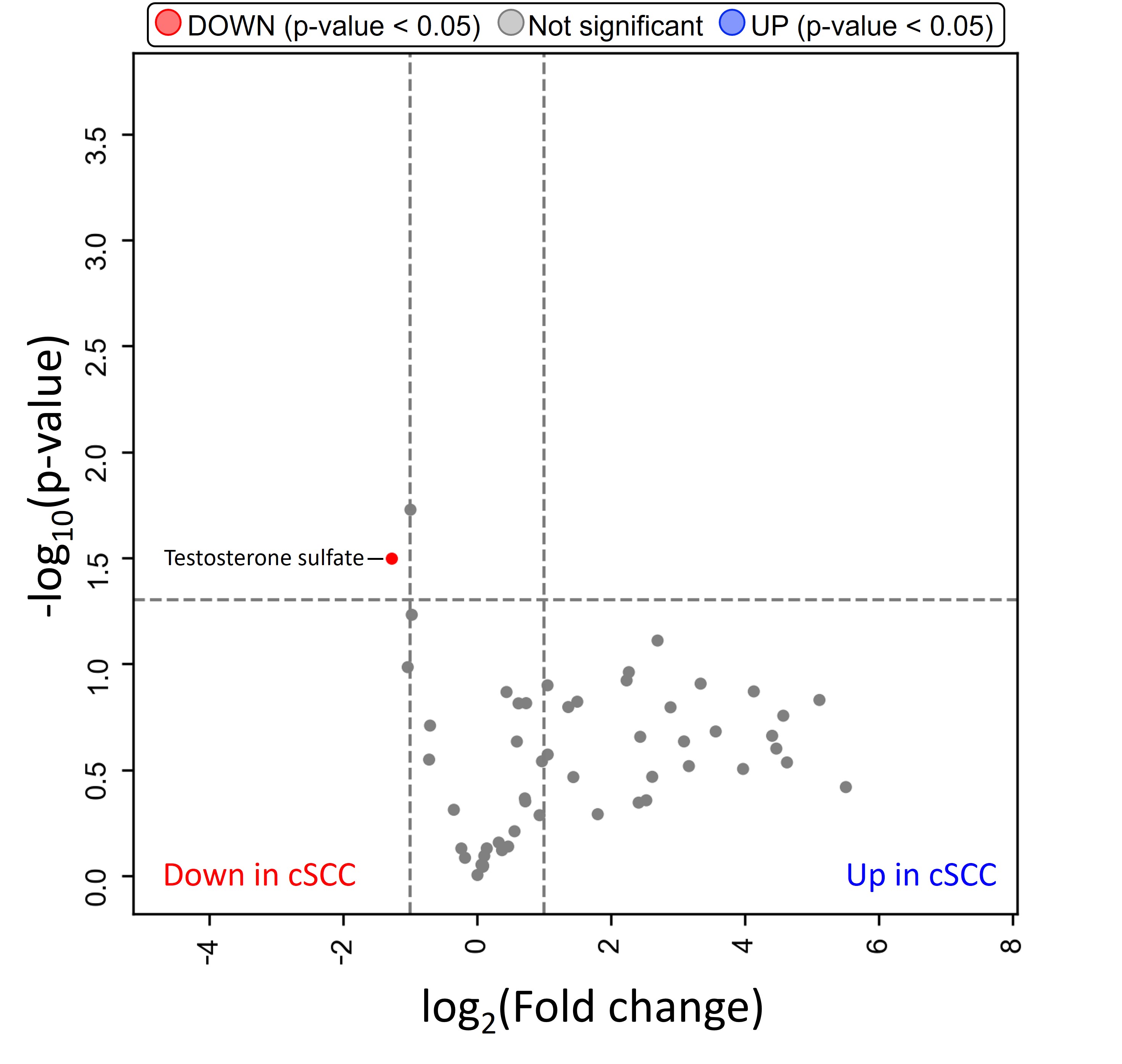

### Fig. S4

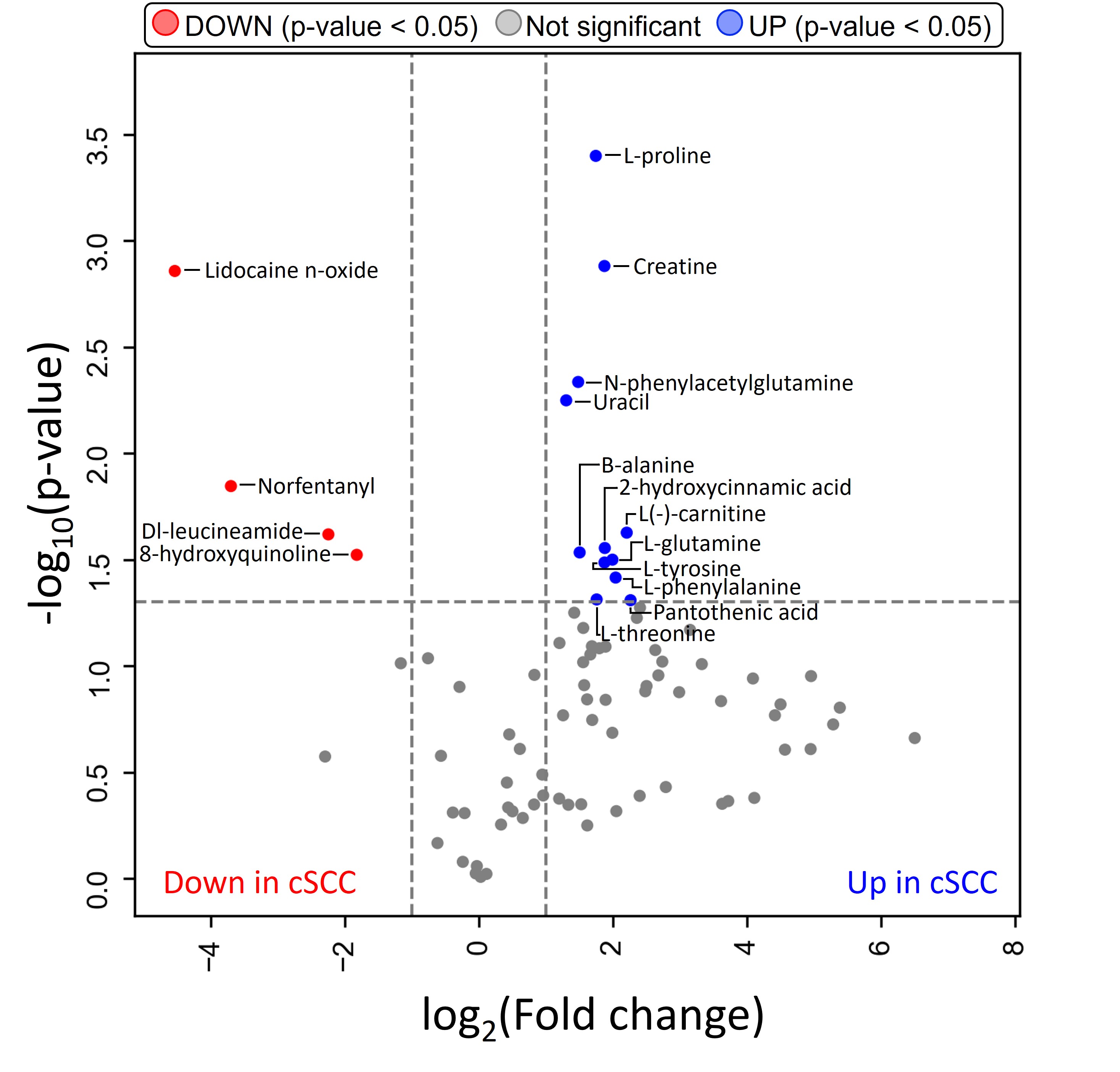

### Fig. S5

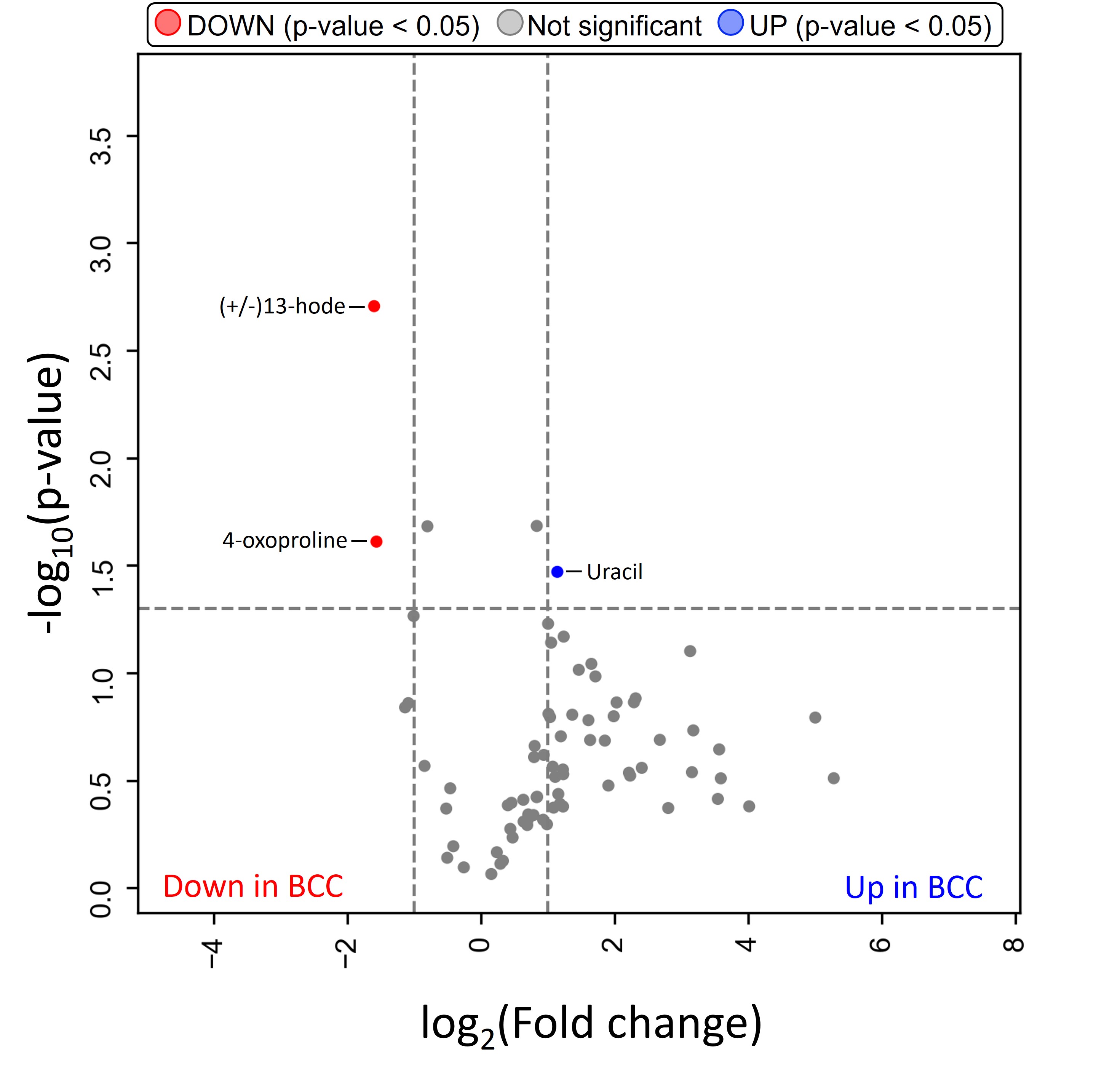

### Fig. S6

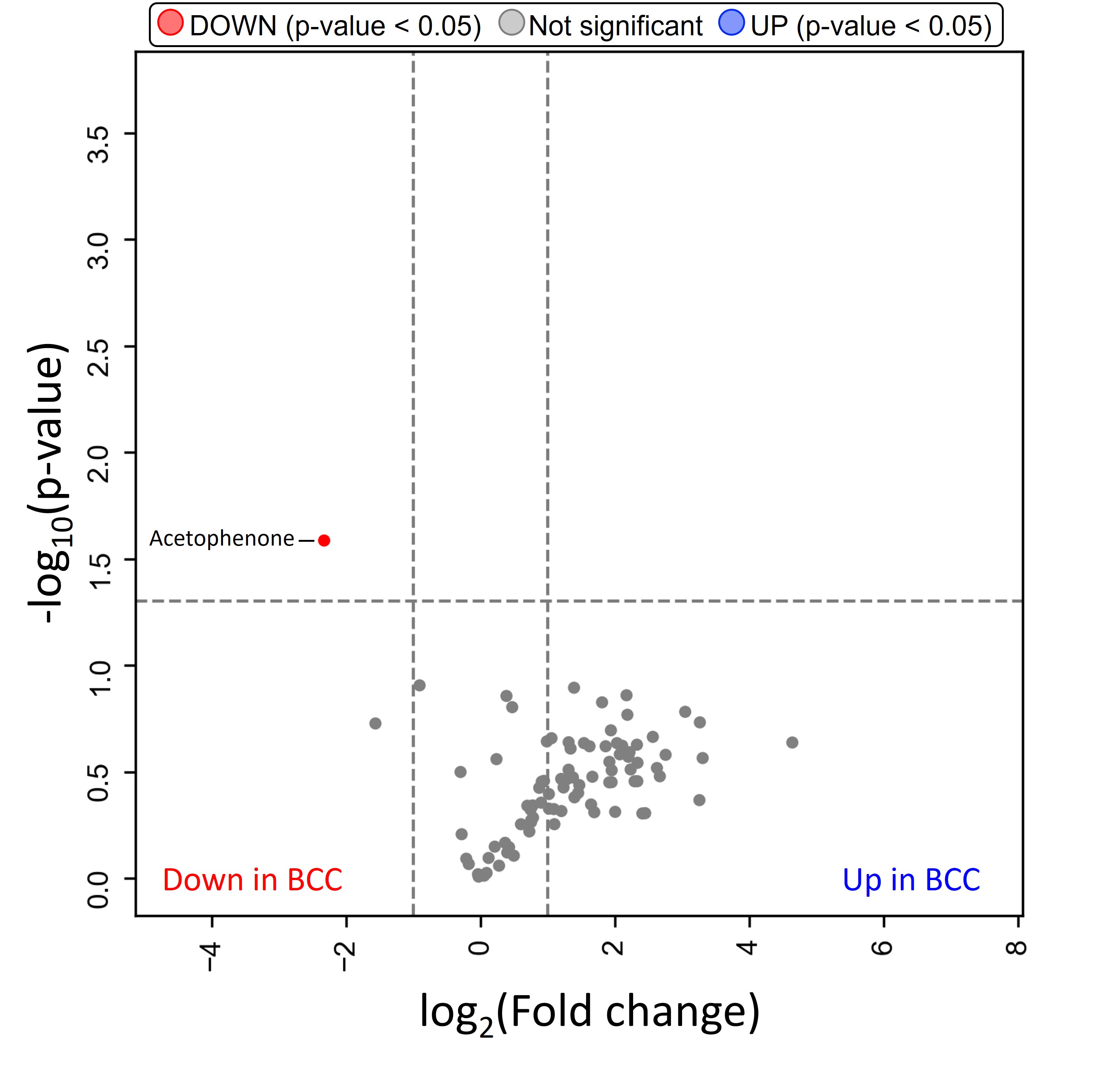

### Fig. S7

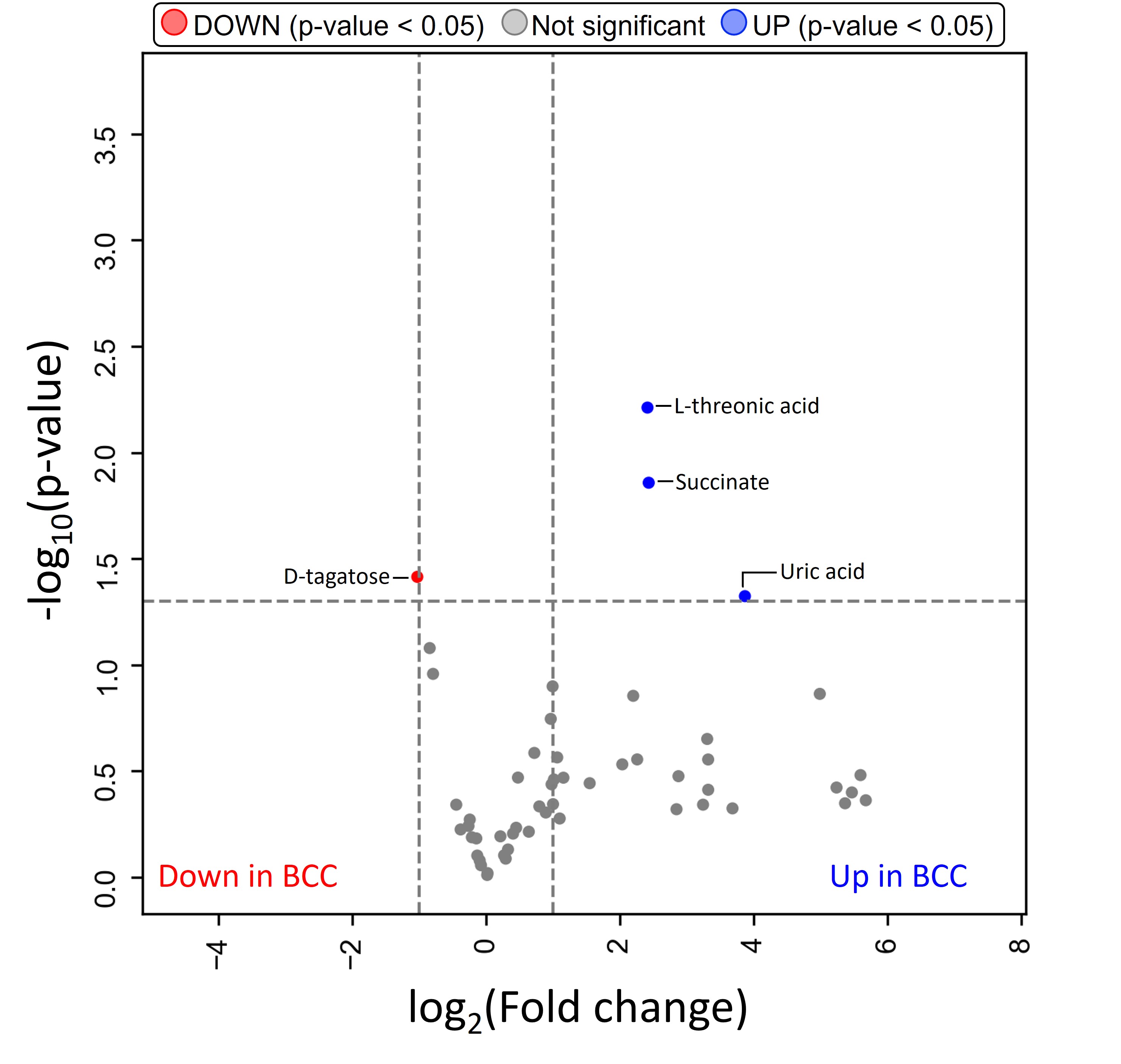

### Fig. S8

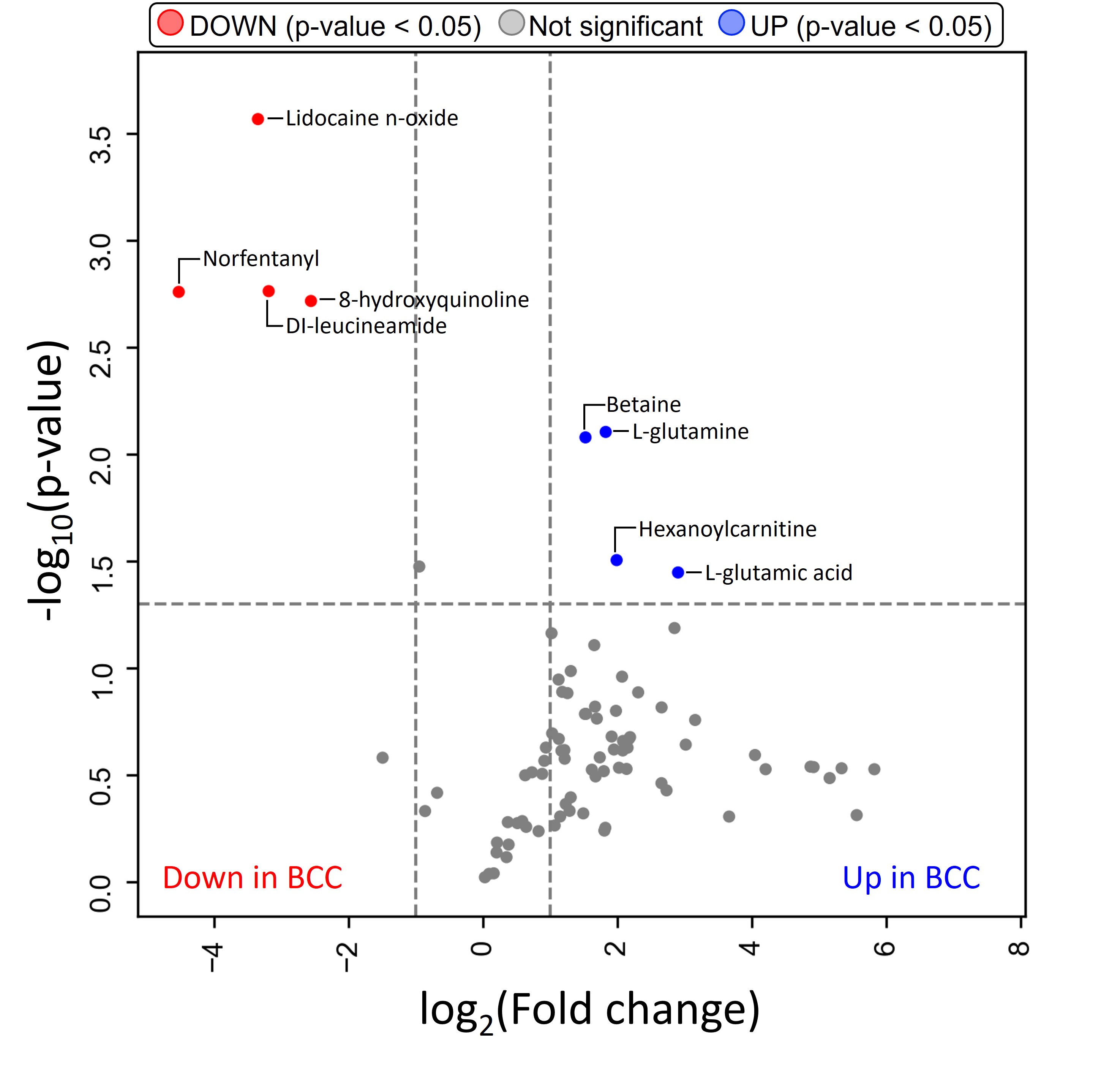

### Fig. S9

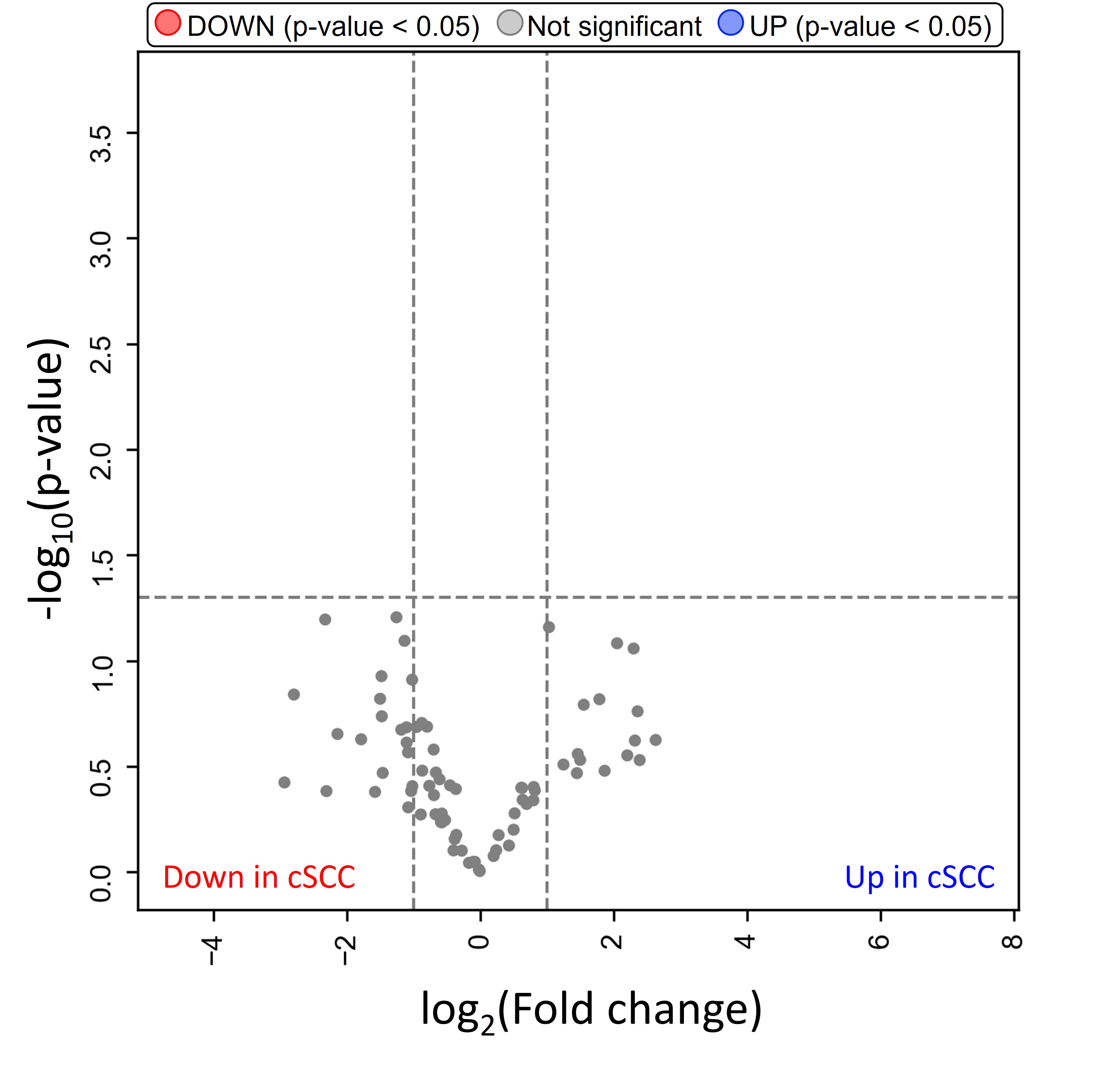

### Fig. S10

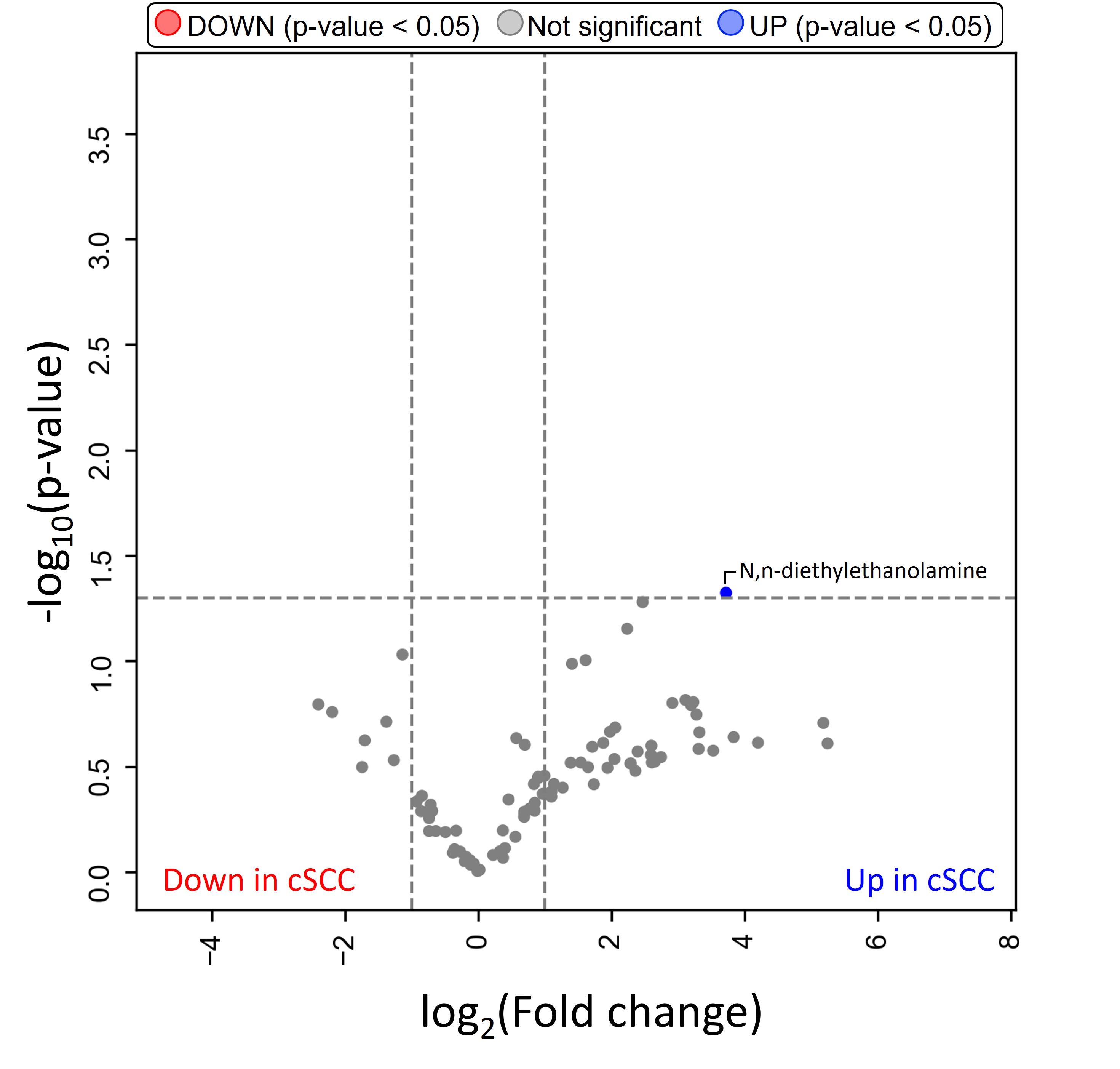

### Fig. S11

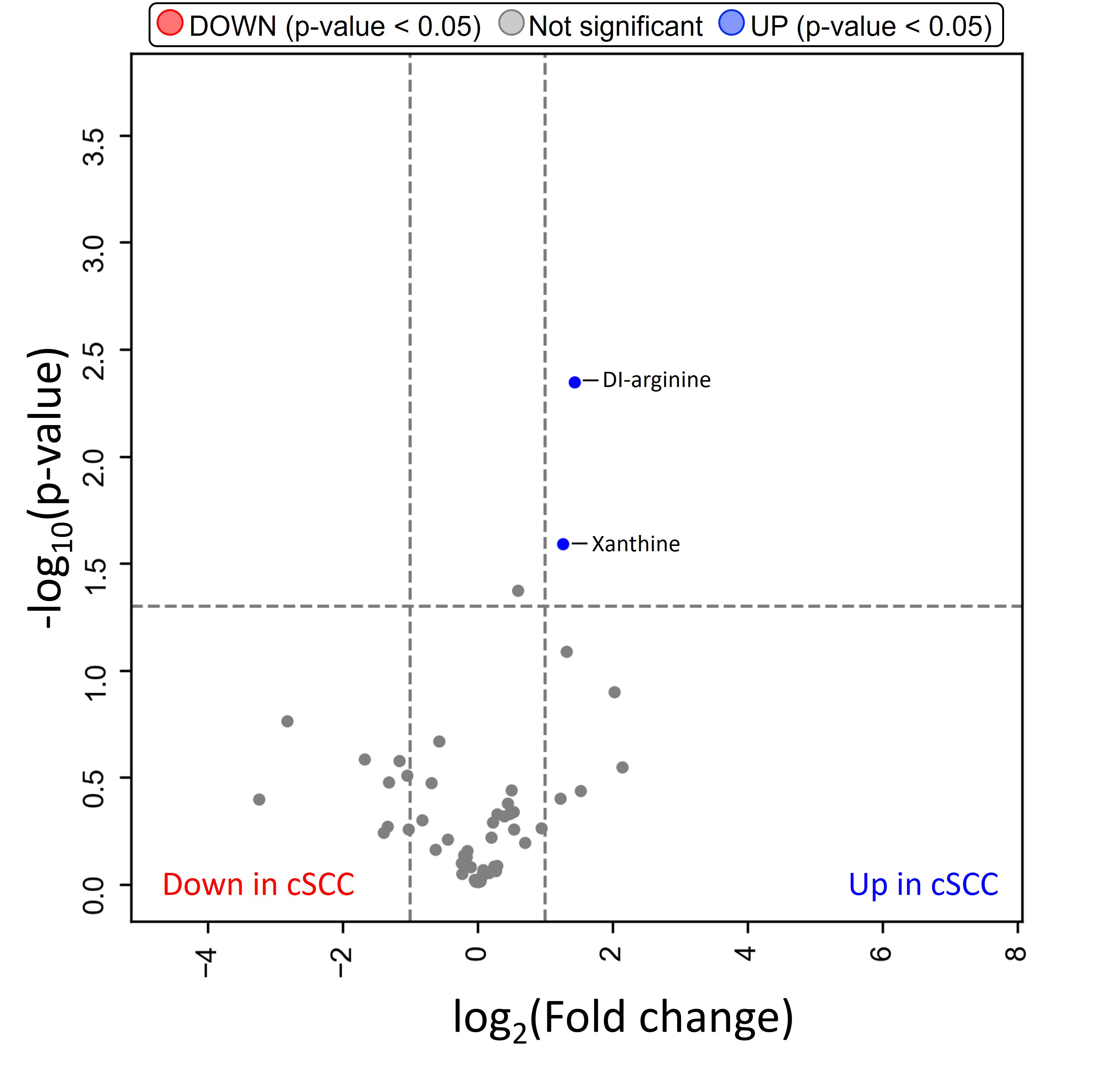

### Fig. S12

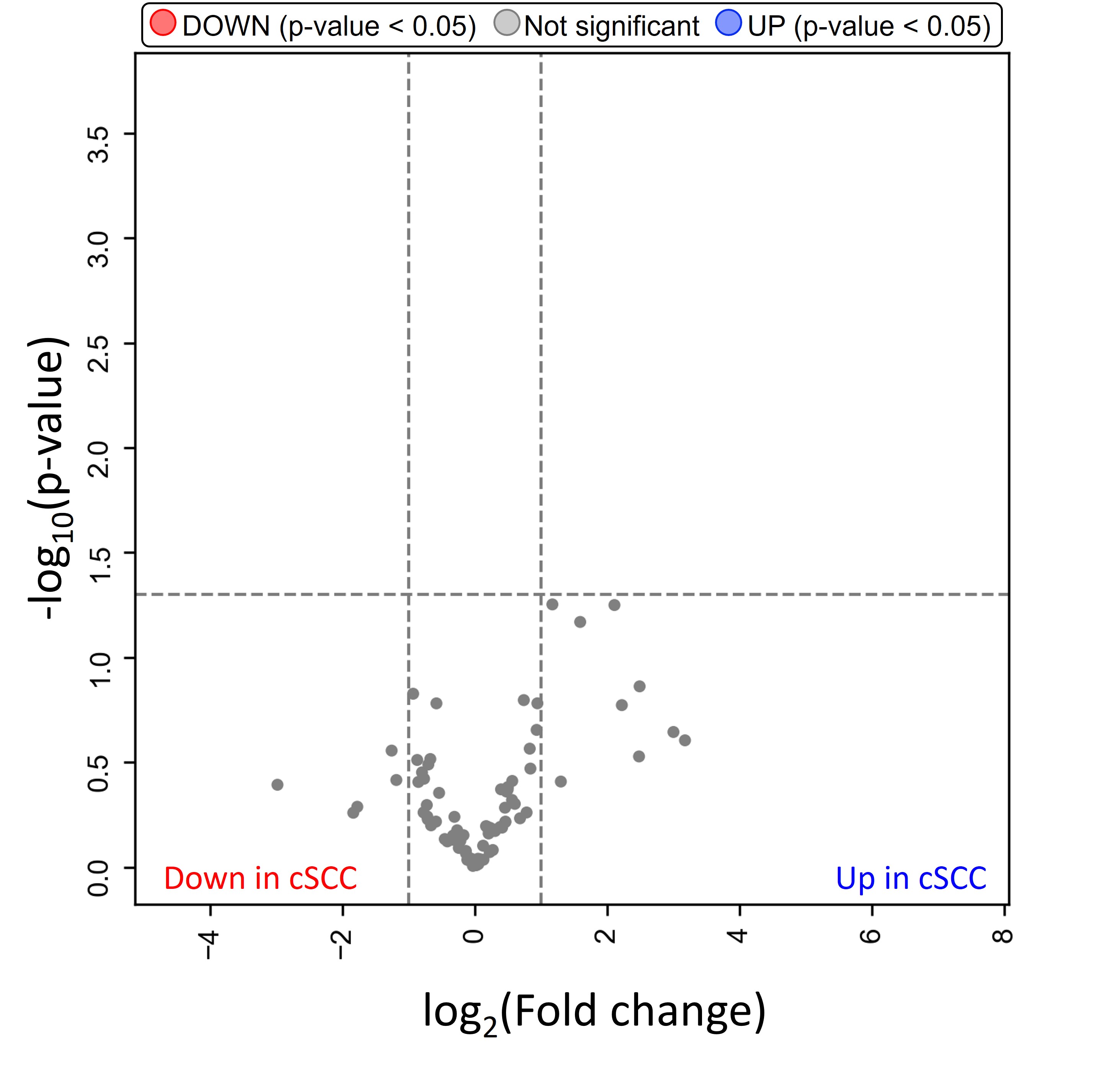
